# Effects of 105 biological, socioeconomic, behavioural, and environmental factors on the risk of SARS-CoV-2 infection and a severe course of Covid-19: A prospective longitudinal study

**DOI:** 10.1101/2021.08.31.21262906

**Authors:** Jaroslav Flegr, Pavel Flegr, Lenka Příplatová

## Abstract

The confirmed number of SARS-CoV-2 infections up to 30 August 2021 is 217 mil. worldwide but information about factors affecting the probability of infection or of a severe course of Covid-19 remains insufficient and often speculative. Only a small number of factors have been rigorously examined, mostly by retrospective or cross-sectional studies. We ran a preregistered study on 5,164 internet users who shared with us information about their exposure to 105 risk factors and reported being Covid negative before the beginning of the fourth wave of Covid-19 in the Czech Republic. After the fourth wave, in which 709 (13.7%) of participants were infected, we used a partial Kendall test controlled for sex, age, and urbanisation to compare the risk of infection and of a severe course of the disease in subjects who originally did and did not report exposure to particular risk factors. After the correction for multiple tests, we identified 13 factors – including male sex, lower age, blood group B, and the larger household size – that increased the risk of infection and 16 factors – including mask wearing, borreliosis in the past, use of vitamin D supplements, or rooibos drinking – that decreased it. We also identified 23 factors that increased the risk of a severe course of Covid-19 and 12 factors that decreased the risk.

## Introduction

According to Covid-19 Data Explorer, up to 29 August, SARS-CoV-2 had infected 217 million of subjects in all continents and has been associated with the death of 4.51 million of persons. Despite the exceptional impact of the Covid-19 pandemic on public health and world economy, a surprisingly small number of studies has been published about risk factors for SARS-CoV-19 infection and about factors that protect individuals against the infection. Search in bibliographic databases for ‘risk factor’ AND ‘Covid’ resulted in 1,583 hits in WOS and 1,853 hits in PubMed (as of 23 July 2021) but an overwhelming majority of original articles reported only risk factors for a severe course of the disease or death in the population of Covid-19 patients, and nearly all articles that dealt with the general population focused on the risk of a severe course or death of Covid-19. Studies searching for risk factors of any (both symptomatic and asymptomatic) infection are surprisingly rare. All in all, less than twenty papers presented the results of prospective longitudinal studies on the risk (or protective) factors associated with Covid-19.

Moreover, the range of factors examined by these retrospective or cross-sectional studies was rather limited. Factors significantly or non-significantly associated with Covid-19 were sex ^1^, age ^1^, ethnicity ^2^, urbanisation ^3^, residence in a multifamily unit ^4^, BMI and obesity ^2,5,6^, smoking ^2,7^, physical fitness and forced expiratory volume ^2^, the number of daily contacts ^3^, wearing masks and washing hands ^3^, socioeconomic deprivation ^2^, particular AB0 blood groups [8-10], Rh factor [10], vitamin D deficiency [11], high-density lipoprotein level ^2^, use of immunosuppressants ^8^, and a growing set of comorbidities – cardiovascular disease, chronic obstructive pulmonary disease (COPD), chronic kidney disease, dementia, hypertension and functional dependence ^6^, and toxoplasmosis ^9^. Other factors, such as contact with animals, have been suggested only on a theoretical basis ^10^ or are merely discussed in non-scientific sources, such as popular literature or the internet. The main aim of the present exploratory study was to perform a systematic investigation of both the known and still unknown factors which might positively or negatively affect the risk of SARS-CoV-2 infection, and to search for factors that might affect the risk of a severe course of Covid-19. To this purpose, we ran a large prospective longitudinal study on the 5,164 originally Covid-negative subjects. To avoid possible cherry-picking artifacts, we preregistered the study before the start of data collection, reported the results of all – both significant and non-significant – tests, and controlled for the effect of multiple statistical tests by the Benjamini-Hochberg procedure.

## Results

In total, 8,084 subjects completed both questionnaires. We excluded 827 subjects who finished the first questionnaire in under 300 seconds or the second questionnaire in under 600 seconds and those who were younger than 15 years. From the remaining 7,257, we excluded 1,262 (17.4%) subjects who had been diagnosed with Covid-19 already before answering the first questionnaire. From the remaining 5,995 subjects, we excluded 578 participants who had not been diagnosed with Covid-19 but suspected they had suffered from it, 13 subjects who were awaiting the results of diagnostic tests when filling the second questionnaire, and 240 subjects who did not respond to the question about their infection status. The final set of originally Covid-negative subjects thus consisted of 5,164 responders: 1,746 men (mean age 42.10, SD 12.28), 3,411 women (mean age 43.46, SD 11.96), and 7 subjects who did not answer the question about their sex (they were included only in tests of the whole population, and only age and urbanisation were controlled for in partial Kendall tests). The difference in age between men and women was significant (t_3437_ = −3.82, p = 0.0001). This set contained 709 (13.7%) subjects who did and 4,455 (86.3%) who did not contract a SARS-CoV-2 infection between completing the first and the second questionnaire. The incidence of infected individuals was non-significantly lower in the 3,411 women (12.13%) than in the 1,746 men (13.52%) (OR = 0.889, C.I._95_ = 0.751-1.054, Chi^2^ = 1.82, p = 0.177); the effect of sex did, however, turn significant when the more sensitive partial Kendall correlation test controlled for age and urbanisation was applied (see Table 3). Other characteristics of the population are described in Tables 1 and 2. The average time since the start of Covid-19 infection was 69.7 days. The mental health of women (p = 1.1 10^−6^) and physical health of both men (p = 5.0 10^−45^) and women (p = 2.6 10^−119^) who had had a Covid-19 infection was significantly worse than in those who avoided the infection (see Fig. 1).

**Table 1.** Exposure to various factors in Covid-negative and Covid-positive subjects The table shows the counts (columns 2–5) and corresponding percentages (columns 6–9) of subjects who had not and those who had been exposed to factors listed in column 1 in subjects who were and those who were not diagnosed with Covid-19, odds ratio (OR), and 95% confidence intervals for the OR.

|  | Number of cases |  |  |  | Fraction of cases |  |  |  | OR | CI<sub>95</sub><br>Low | CI<sub>95</sub><br>High |
| --- | --- | --- | --- | --- | --- | --- | --- | --- | --- | --- | --- |
|  | Covid-minus |  | Covid-plus |  | Covid-minus |  | Covid-plus |  |  |  |  |
|  | No | Yes | No | Yes | No | Yes | No | Yes |  |  |  |
| Female sex | 1,490 | 2,959 | 256 | 452 | 33.5 | 66.5 | 36.2 | 63.8 | 0.89 | 0.75 | 1.05 |
| Blood group A | 2,067 | 1,200 | 335 | 192 | 63.3 | 36.7 | 63.6 | 36.4 | 0.99 | 0.81 | 1.20 |
| Blood group B | 2,614 | 653 | 403 | 124 | 80.0 | 20.0 | 76.5 | 23.5 | 1.23 | 0.98 | 1.54 |
| Blood group AB | 2,979 | 288 | 482 | 45 | 91.2 | 8.8 | 91.5 | 8.5 | 0.97 | 0.68 | 1.35 |
| Blood group 0 | 2,141 | 1,126 | 361 | 166 | 65.5 | 34.5 | 68.5 | 31.5 | 0.87 | 0.71 | 1.07 |
| Rh-positivity | 667 | 2,376 | 102 | 390 | 21.9 | 78.1 | 20.7 | 79.3 | 1.07 | 0.85 | 1.37 |
| Rh-heterozygosity | 667 | 256 | 102 | 37 | 72.3 | 27.7 | 73.4 | 26.6 | 0.95 | 0.61 | 1.43 |
| Living single | 3,734 | 713 | 623 | 85 | 84.0 | 16.0 | 88.0 | 12.0 | 0.71 | 0.56 | 0.91 |
| Wearing glasses | 1,833 | 1,177 | 257 | 168 | 60.9 | 39.1 | 60.5 | 39.5 | 1.02 | 0.82 | 1.26 |
| Tobacco smoking | 2,664 | 683 | 413 | 79 | 79.6 | 20.4 | 83.9 | 16.1 | 0.75 | 0.57 | 0.97 |
| Marihuana consumption | 3,223 | 124 | 478 | 14 | 96.3 | 3.7 | 97.2 | 2.8 | 0.76 | 0.40 | 1.34 |
| Daily alcohol consumption | 3,042 | 305 | 450 | 42 | 90.9 | 9.1 | 91.5 | 8.5 | 0.93 | 0.65 | 1.31 |
| Snoring | 2,690 | 657 | 396 | 96 | 80.4 | 19.6 | 80.5 | 19.5 | 0.99 | 0.77 | 1.27 |
| Frequent singing | 2,969 | 378 | 424 | 68 | 88.7 | 11.3 | 86.2 | 13.8 | 1.26 | 0.94 | 1.67 |
| Sport | 2,326 | 1,021 | 319 | 173 | 69.5 | 30.5 | 64.8 | 35.2 | 1.24 | 1.01 | 1.51 |
| Cold water swimming | 2,995 | 352 | 422 | 70 | 89.5 | 10.5 | 85.8 | 14.2 | 1.41 | 1.05 | 1.87 |
| Vitamins and supplements | 1,217 | 2130 | 203 | 289 | 36.4 | 63.6 | 41.3 | 58.7 | 0.81 | 0.67 | 0.99 |
| Volunteering | 3,136 | 211 | 459 | 33 | 93.7 | 6.3 | 93.3 | 6.7 | 1.07 | 0.71 | 1.57 |
| Walking in nature | 1,070 | 1,023 | 133 | 112 | 51.1 | 48.9 | 54.3 | 45.7 | 0.88 | 0.67 | 1.16 |
| Frequent use of sauna | 1,925 | 168 | 228 | 17 | 92.0 | 8.0 | 93.1 | 6.9 | 0.85 | 0.48 | 1.44 |
| Dog | 1,921 | 2,518 | 299 | 409 | 43.3 | 56.7 | 42.2 | 57.8 | 1.04 | 0.89 | 1.23 |
| Cat | 2,243 | 2,196 | 350 | 358 | 50.5 | 49.5 | 49.4 | 50.6 | 1.04 | 0.89 | 1.23 |
| Bird | 3,597 | 842 | 572 | 136 | 81.0 | 19.0 | 80.8 | 19.2 | 1.02 | 0.82 | 1.25 |
| Reptile | 4,116 | 323 | 646 | 62 | 92.7 | 7.3 | 91.2 | 8.8 | 1.22 | 0.90 | 1.63 |
| Fish | 3,240 | 1,199 | 524 | 184 | 73.0 | 27.0 | 74.0 | 26.0 | 0.95 | 0.79 | 1.14 |
| Rabbit | 3,656 | 783 | 576 | 132 | 82.4 | 17.6 | 81.4 | 18.6 | 1.07 | 0.87 | 1.32 |
| Guinea pigs, hamster | 3,159 | 1,280 | 486 | 222 | 71.2 | 28.8 | 68.6 | 31.4 | 1.13 | 0.95 | 1.34 |
| Fowls | 3,723 | 716 | 591 | 117 | 83.9 | 16.1 | 83.5 | 16.5 | 1.03 | 0.82 | 1.28 |
| Goats, sheep | 4,239 | 200 | 683 | 25 | 95.5 | 4.5 | 96.5 | 3.5 | 0.78 | 0.49 | 1.19 |
| Mouse, rat | 3,027 | 311 | 434 | 58 | 90.7 | 9.3 | 88.2 | 11.8 | 1.30 | 0.95 | 1.76 |
| Pig | 3,193 | 145 | 469 | 23 | 95.7 | 4.3 | 95.3 | 4.7 | 1.08 | 0.66 | 1.71 |
| Horse | 3,241 | 97 | 473 | 19 | 97.1 | 2.9 | 96.1 | 3.9 | 1.34 | 0.77 | 2.24 |
| Being overweight | 2,064 | 1,283 | 303 | 189 | 61.7 | 38.3 | 61.6 | 38.4 | 1.00 | 0.82 | 1.22 |
| Overweight BMI>25 | 1,877 | 2,438 | 292 | 399 | 43.5 | 56.5 | 42.3 | 57.7 | 1.05 | 0.89 | 1.24 |
| Obesity BMI>30 | 3,289 | 1,026 | 517 | 174 | 76.2 | 23.8 | 74.8 | 25.2 | 1.08 | 0.89 | 1.30 |
| Underweight | 4,221 | 94 | 676 | 15 | 97.8 | 2.2 | 97.8 | 2.2 | 1.00 | 0.53 | 1.74 |
| Diabetes | 3,215 | 132 | 477 | 15 | 96.1 | 3.9 | 97.0 | 3.0 | 0.77 | 0.41 | 1.32 |
| Cardiovascular problems | 3,044 | 303 | 454 | 38 | 90.9 | 9.1 | 92.3 | 7.7 | 0.84 | 0.58 | 1.20 |
| Asthma | 2,998 | 349 | 441 | 51 | 89.6 | 10.4 | 89.6 | 10.4 | 0.99 | 0.71 | 1.36 |
| Chronic obstructive pulmonary disease | 3,272 | 75 | 486 | 6 | 97.8 | 2.2 | 98.8 | 1.2 | 0.54 | 0.19 | 1.24 |
| Immunodeficiency | 3,037 | 310 | 454 | 38 | 90.7 | 9.3 | 92.3 | 7.7 | 0.82 | 0.56 | 1.17 |
| Allergy | 2,521 | 826 | 365 | 127 | 75.3 | 24.7 | 74.2 | 25.8 | 1.06 | 0.85 | 1.32 |
| Autoimmunity | 1,877 | 216 | 215 | 30 | 89.7 | 10.3 | 87.8 | 12.2 | 1.21 | 0.78 | 1.84 |
| Toxoplasmosis | 633 | 153 | 104 | 25 | 80.5 | 19.5 | 80.6 | 19.4 | 0.99 | 0.59 | 1.62 |
| Borreliosis | 993 | 560 | 179 | 63 | 63.9 | 36.1 | 74.0 | 26.0 | 0.62 | 0.45 | 0.85 |
| Depression | 2,978 | 369 | 437 | 55 | 89.0 | 11.0 | 88.8 | 11.2 | 1.02 | 0.74 | 1.38 |
| Anxiety | 2,588 | 759 | 361 | 131 | 77.3 | 22.7 | 73.4 | 26.6 | 1.24 | 0.99 | 1.54 |
| Vitamin A | 1,026 | 93 | 83 | 7 | 91.7 | 8.3 | 92.2 | 7.8 | 0.93 | 0.35 | 2.08 |
| Vitamin B | 670 | 449 | 52 | 38 | 59.9 | 40.1 | 57.8 | 42.2 | 1.09 | 0.69 | 1.72 |
| Vitamin C | 367 | 752 | 26 | 64 | 32.8 | 67.2 | 28.9 | 71.1 | 1.20 | 0.74 | 2.01 |
| Vitamin D | 328 | 791 | 36 | 54 | 29.3 | 70.7 | 40.0 | 60.0 | 0.62 | 0.39 | 1.00 |
| Vitamin E | 990 | 129 | 82 | 8 | 88.5 | 11.5 | 91.1 | 8.9 | 0.75 | 0.31 | 1.59 |
| Vitamin K | 1,024 | 95 | 83 | 7 | 91.5 | 8.5 | 92.2 | 7.8 | 0.91 | 0.34 | 2.04 |
| Magnesium | 644 | 475 | 52 | 38 | 57.6 | 42.4 | 57.8 | 42.2 | 0.99 | 0.62 | 1.56 |
| Zinc | 792 | 327 | 69 | 21 | 70.8 | 29.2 | 76.7 | 23.3 | 0.74 | 0.42 | 1.24 |
| Selenium fluorine iodine | 1,017 | 102 | 82 | 8 | 90.9 | 9.1 | 91.1 | 8.9 | 0.97 | 0.40 | 2.09 |
| Calcium | 920 | 199 | 76 | 14 | 82.2 | 17.8 | 84.4 | 15.6 | 0.85 | 0.44 | 1.56 |
| Iron | 983 | 136 | 82 | 8 | 87.8 | 12.2 | 91.1 | 8.9 | 0.71 | 0.29 | 1.50 |
| Antioxidants | 994 | 125 | 78 | 12 | 88.8 | 11.2 | 86.7 | 13.3 | 1.22 | 0.59 | 2.34 |
| Fatty acids | 841 | 278 | 66 | 24 | 75.2 | 24.8 | 73.3 | 26.7 | 1.10 | 0.65 | 1.82 |
| Coenzyme Q10 | 1,055 | 64 | 85 | 5 | 94.3 | 5.7 | 94.4 | 5.6 | 0.97 | 0.30 | 2.48 |
| Apple cider vinegar | 1,038 | 81 | 83 | 7 | 92.8 | 7.2 | 92.2 | 7.8 | 1.08 | 0.41 | 2.43 |
| Coconut oil | 1,033 | 86 | 84 | 6 | 92.3 | 7.7 | 93.3 | 6.7 | 0.86 | 0.30 | 2.03 |
| Echinacea | 1,028 | 91 | 83 | 7 | 91.9 | 8.1 | 92.2 | 7.8 | 0.95 | 0.36 | 2.14 |
| Immunglucan | 1,058 | 61 | 86 | 4 | 94.5 | 5.5 | 95.6 | 4.4 | 0.81 | 0.21 | 2.25 |
| Lecithin | 1,087 | 32 | 89 | 1 | 97.1 | 2.9 | 98.9 | 1.1 | 0.38 | 0.01 | 2.34 |
| Dimethyl sulfone | 1,114 | 5 | 90 | 0 | 99.6 | 0.4 | 100.0 |  |  |  |  |
| Chlorine dioxide | 1,114 | 5 | 90 | 0 | 99.6 | 0.4 | 100.0 |  |  |  |  |
| Collagen | 997 | 122 | 83 | 7 | 89.1 | 10.9 | 92.2 | 7.8 | 0.69 | 0.26 | 1.53 |
| Green tea, matcha | 928 | 191 | 73 | 17 | 82.9 | 17.1 | 81.1 | 18.9 | 1.13 | 0.61 | 1.99 |
| Chlorella | 1,066 | 53 | 84 | 6 | 95.3 | 4.7 | 93.3 | 6.7 | 1.44 | 0.49 | 3.47 |
| Ginseng | 1,081 | 38 | 88 | 2 | 96.6 | 3.4 | 97.8 | 2.2 | 0.65 | 0.07 | 2.58 |
| Rooibos | 998 | 121 | 87 | 3 | 89.2 | 10.8 | 96.7 | 3.3 | 0.28 | 0.06 | 0.88 |
| Suppl. for pregnant women | 1,072 | 47 | 88 | 2 | 95.8 | 4.2 | 97.8 | 2.2 | 0.52 | 0.06 | 2.04 |
| Sports supplements | 1,072 | 47 | 87 | 3 | 95.8 | 4.2 | 96.7 | 3.3 | 0.79 | 0.15 | 2.52 |
| Weight loss supplements | 1,109 | 10 | 89 | 1 | 99.1 | 0.9 | 98.9 | 1.1 | 1.25 | 0.03 | 8.94 |
| Yucca | 1,116 | 3 | 90 | 0 | 99.7 | 0.3 | 100.0 |  |  |  |  |
| Vilcacora | 1,113 | 6 | 90 | 0 | 99.5 | 0.5 | 100.0 |  |  |  |  |
| Lapacho | 1,111 | 8 | 88 | 2 | 99.3 | 0.7 | 97.8 | 2.2 | 3.15 | 0.32 | 16.12 |
| Chinese herbs | 1,095 | 24 | 88 | 2 | 97.9 | 2.1 | 97.8 | 2.2 | 1.04 | 0.12 | 4.29 |
| Medical herbs | 830 | 289 | 62 | 28 | 74.2 | 25.8 | 68.9 | 31.1 | 1.30 | 0.78 | 2.10 |
| Vironal | 1,103 | 16 | 88 | 2 | 98.6 | 1.4 | 97.8 | 2.2 | 1.57 | 0.17 | 6.83 |
| Melatonin | 1,079 | 40 | 84 | 6 | 96.4 | 3.6 | 93.3 | 6.7 | 1.93 | 0.65 | 4.75 |
| Cannabis | 1,065 | 54 | 86 | 4 | 95.2 | 4.8 | 95.6 | 4.4 | 0.92 | 0.24 | 2.58 |
| Aloe vera | 1,072 | 47 | 88 | 2 | 95.8 | 4.2 | 97.8 | 2.2 | 0.52 | 0.06 | 2.04 |
| Homeopathic | 1,078 | 41 | 84 | 6 | 96.3 | 3.7 | 93.3 | 6.7 | 1.88 | 0.63 | 4.62 |
| Adaptogenic fungi | 1,093 | 26 | 90 | 0 | 97.7 | 2.3 | 100.0 |  |  |  |  |
| Enzymes | 1,111 | 8 | 89 | 1 | 99.3 | 0.7 | 98.9 | 1.1 | 1.56 | 0.03 | 11.86 |
| Flavonoids | 1,100 | 19 | 89 | 1 | 98.3 | 1.7 | 98.9 | 1.1 | 0.65 | 0.02 | 4.19 |
| Sea buckthorn | 964 | 155 | 78 | 12 | 86.1 | 13.9 | 86.7 | 13.3 | 0.96 | 0.46 | 1.82 |
| Supplements other | 993 | 126 | 80 | 10 | 88.7 | 11.3 | 88.9 | 11.1 | 0.99 | 0.44 | 1.97 |

**Table 2.**
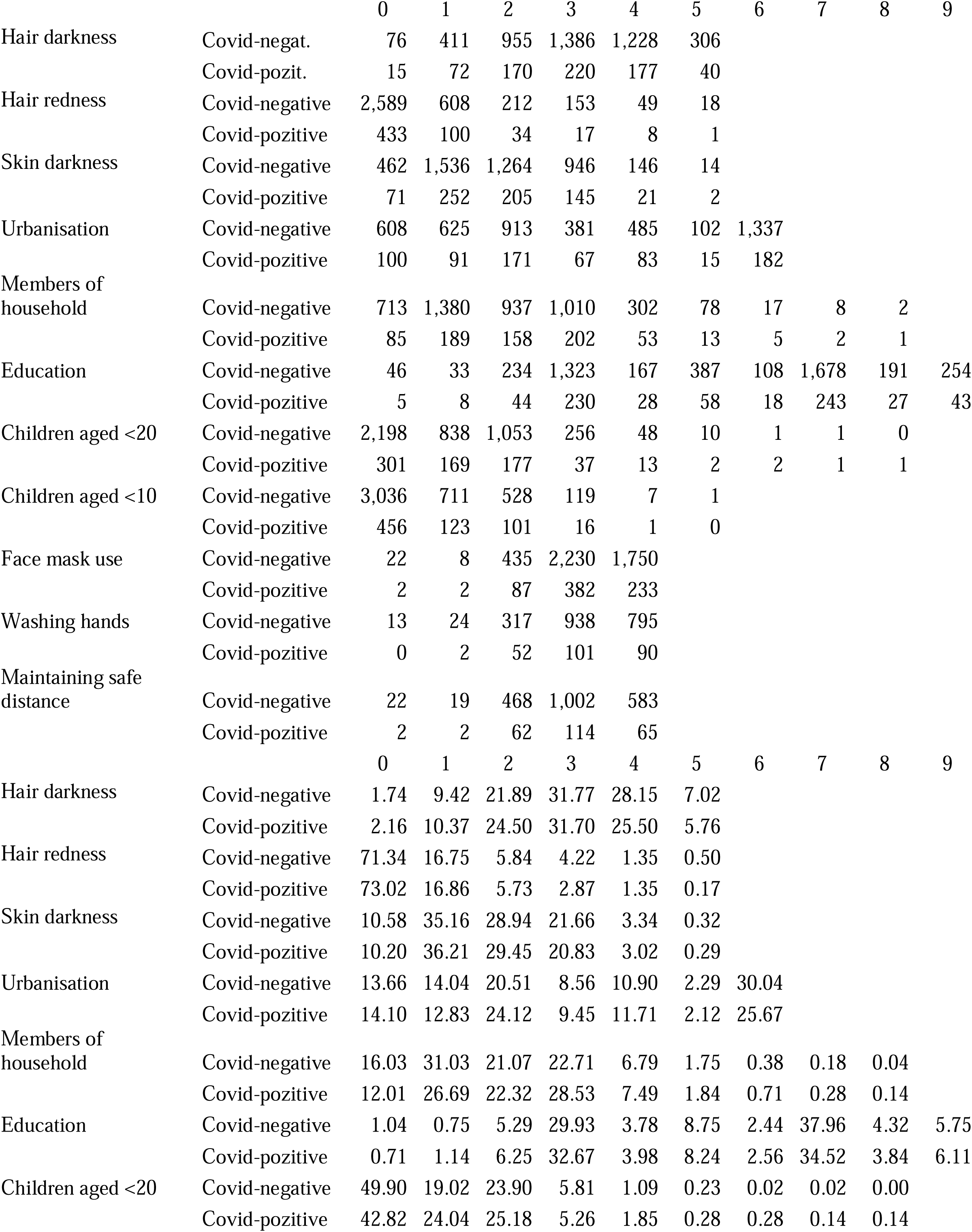
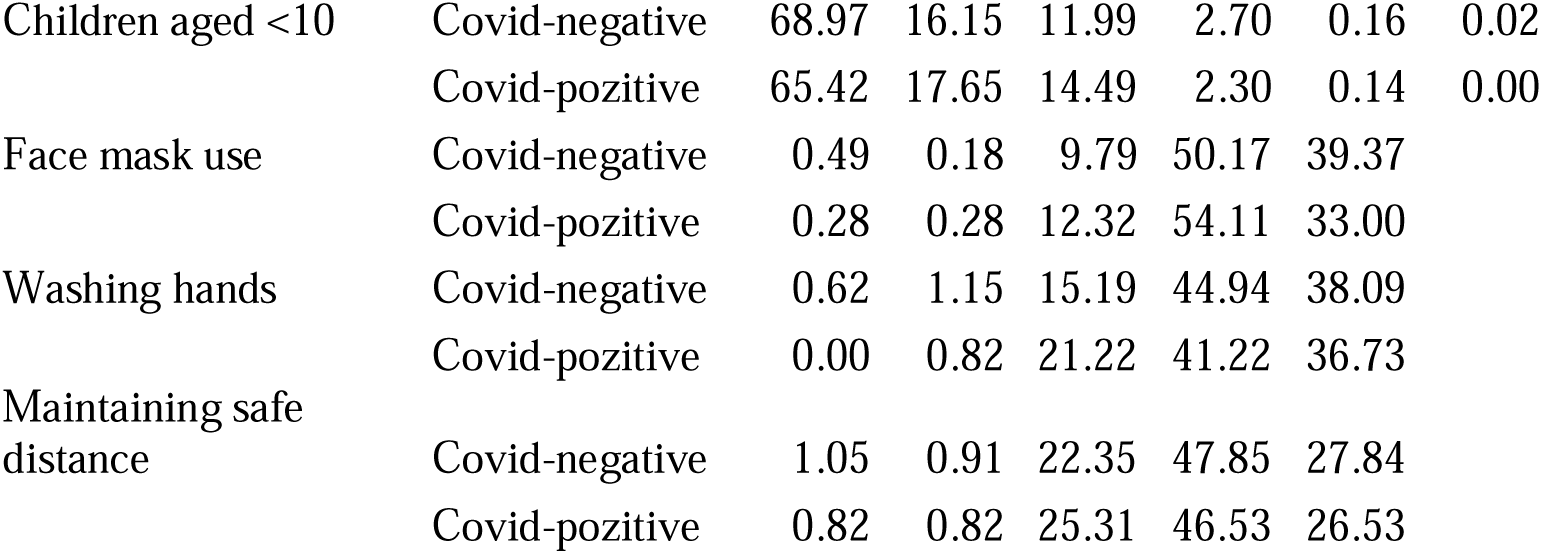
Distributions of responses to the questions with ordinal scale in individuals who had and had not been diagnosed with Covid-19. The first part of the table shows the numbers of subjects providing a particular response; the second shows the corresponding percentages. For the meaning of particular response codes, see Material and Methods.

|  |  | 0 | 1 | 2 | 3 | 4 | 5 | 6 | 7 | 8 | 9 |
| --- | --- | --- | --- | --- | --- | --- | --- | --- | --- | --- | --- |
| Hair darkness | Covid-negat. | 76 | 411 | 955 | 1,386 | 1,228 | 306 |  |  |  |  |
|  | Covid-pozit. | 15 | 72 | 170 | 220 | 177 | 40 |  |  |  |  |
| Hair redness | Covid-negative | 2,589 | 608 | 212 | 153 | 49 | 18 |  |  |  |  |
|  | Covid-pozitive | 433 | 100 | 34 | 17 | 8 | 1 |  |  |  |  |
| Skin darkness | Covid-negative | 462 | 1,536 | 1,264 | 946 | 146 | 14 |  |  |  |  |
|  | Covid-pozitive | 71 | 252 | 205 | 145 | 21 | 2 |  |  |  |  |
| Urbanisation | Covid-negative | 608 | 625 | 913 | 381 | 485 | 102 | 1,337 |  |  |  |
|  | Covid-pozitive | 100 | 91 | 171 | 67 | 83 | 15 | 182 |  |  |  |
| Members of household | Covid-negative | 713 | 1,380 | 937 | 1,010 | 302 | 78 | 17 | 8 | 2 |  |
|  | Covid-pozitive | 85 | 189 | 158 | 202 | 53 | 13 | 5 | 2 | 1 |  |
| Education | Covid-negative | 46 | 33 | 234 | 1,323 | 167 | 387 | 108 | 1,678 | 191 | 254 |
|  | Covid-pozitive | 5 | 8 | 44 | 230 | 28 | 58 | 18 | 243 | 27 | 43 |
| Children aged <20 | Covid-negative | 2,198 | 838 | 1,053 | 256 | 48 | 10 | 1 | 1 | 0 |  |
|  | Covid-pozitive | 301 | 169 | 177 | 37 | 13 | 2 | 2 | 1 | 1 |  |
| Children aged <10 | Covid-negative | 3,036 | 711 | 528 | 119 | 7 | 1 |  |  |  |  |
|  | Covid-pozitive | 456 | 123 | 101 | 16 | 1 | 0 |  |  |  |  |
| Face mask use | Covid-negative | 22 | 8 | 435 | 2,230 | 1,750 |  |  |  |  |  |
|  | Covid-pozitive | 2 | 2 | 87 | 382 | 233 |  |  |  |  |  |
| Washing hands | Covid-negative | 13 | 24 | 317 | 938 | 795 |  |  |  |  |  |
|  | Covid-pozitive | 0 | 2 | 52 | 101 | 90 |  |  |  |  |  |
| Maintaining safe distance | Covid-negative | 22 | 19 | 468 | 1,002 | 583 |  |  |  |  |  |
|  | Covid-pozitive | 2 | 2 | 62 | 114 | 65 |  |  |  |  |  |
|  |  | 0 | 1 | 2 | 3 | 4 | 5 | 6 | 7 | 8 | 9 |
| Hair darkness | Covid-negative | 1.74 | 9.42 | 21.89 | 31.77 | 28.15 | 7.02 |  |  |  |  |
|  | Covid-pozitive | 2.16 | 10.37 | 24.50 | 31.70 | 25.50 | 5.76 |  |  |  |  |
| Hair redness | Covid-negative | 71.34 | 16.75 | 5.84 | 4.22 | 1.35 | 0.50 |  |  |  |  |
|  | Covid-pozitive | 73.02 | 16.86 | 5.73 | 2.87 | 1.35 | 0.17 |  |  |  |  |
| Skin darkness | Covid-negative | 10.58 | 35.16 | 28.94 | 21.66 | 3.34 | 0.32 |  |  |  |  |
|  | Covid-pozitive | 10.20 | 36.21 | 29.45 | 20.83 | 3.02 | 0.29 |  |  |  |  |
| Urbanisation | Covid-negative | 13.66 | 14.04 | 20.51 | 8.56 | 10.90 | 2.29 | 30.04 |  |  |  |
|  | Covid-pozitive | 14.10 | 12.83 | 24.12 | 9.45 | 11.71 | 2.12 | 25.67 |  |  |  |
| Members of household | Covid-negative | 16.03 | 31.03 | 21.07 | 22.71 | 6.79 | 1.75 | 0.38 | 0.18 | 0.04 |  |
|  | Covid-pozitive | 12.01 | 26.69 | 22.32 | 28.53 | 7.49 | 1.84 | 0.71 | 0.28 | 0.14 |  |
| Education | Covid-negative | 1.04 | 0.75 | 5.29 | 29.93 | 3.78 | 8.75 | 2.44 | 37.96 | 4.32 | 5.75 |
|  | Covid-pozitive | 0.71 | 1.14 | 6.25 | 32.67 | 3.98 | 8.24 | 2.56 | 34.52 | 3.84 | 6.11 |
| Children aged <20 | Covid-negative | 49.90 | 19.02 | 23.90 | 5.81 | 1.09 | 0.23 | 0.02 | 0.02 | 0.00 |  |
|  | Covid-pozitive | 42.82 | 24.04 | 25.18 | 5.26 | 1.85 | 0.28 | 0.28 | 0.14 | 0.14 |  |

|  |  |  |  |  |  |  |  |
| --- | --- | --- | --- | --- | --- | --- | --- |
| Children aged <10 | Covid-negative | 68.97 | 16.15 | 11.99 | 2.70 | 0.16 | 0.02 |
|  | Covid-pozitive | 65.42 | 17.65 | 14.49 | 2.30 | 0.14 | 0.00 |
| Face mask use | Covid-negative | 0.49 | 0.18 | 9.79 | 50.17 | 39.37 |  |
|  | Covid-pozitive | 0.28 | 0.28 | 12.32 | 54.11 | 33.00 |  |
| Washing hands | Covid-negative | 0.62 | 1.15 | 15.19 | 44.94 | 38.09 |  |
|  | Covid-pozitive | 0.00 | 0.82 | 21.22 | 41.22 | 36.73 |  |
| Maintaining safe distance | Covid-negative | 1.05 | 0.91 | 22.35 | 47.85 | 27.84 |  |
|  | Covid-pozitive | 0.82 | 0.82 | 25.31 | 46.53 | 26.53 |  |

**Table 3.**
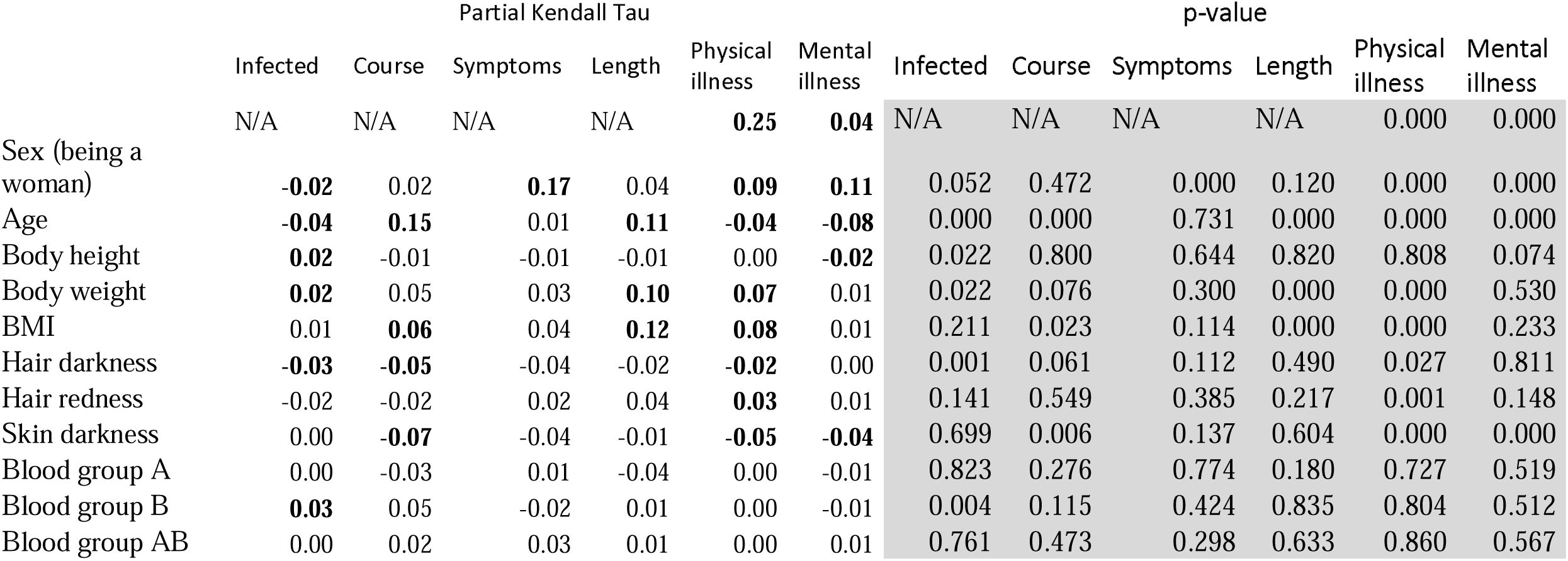
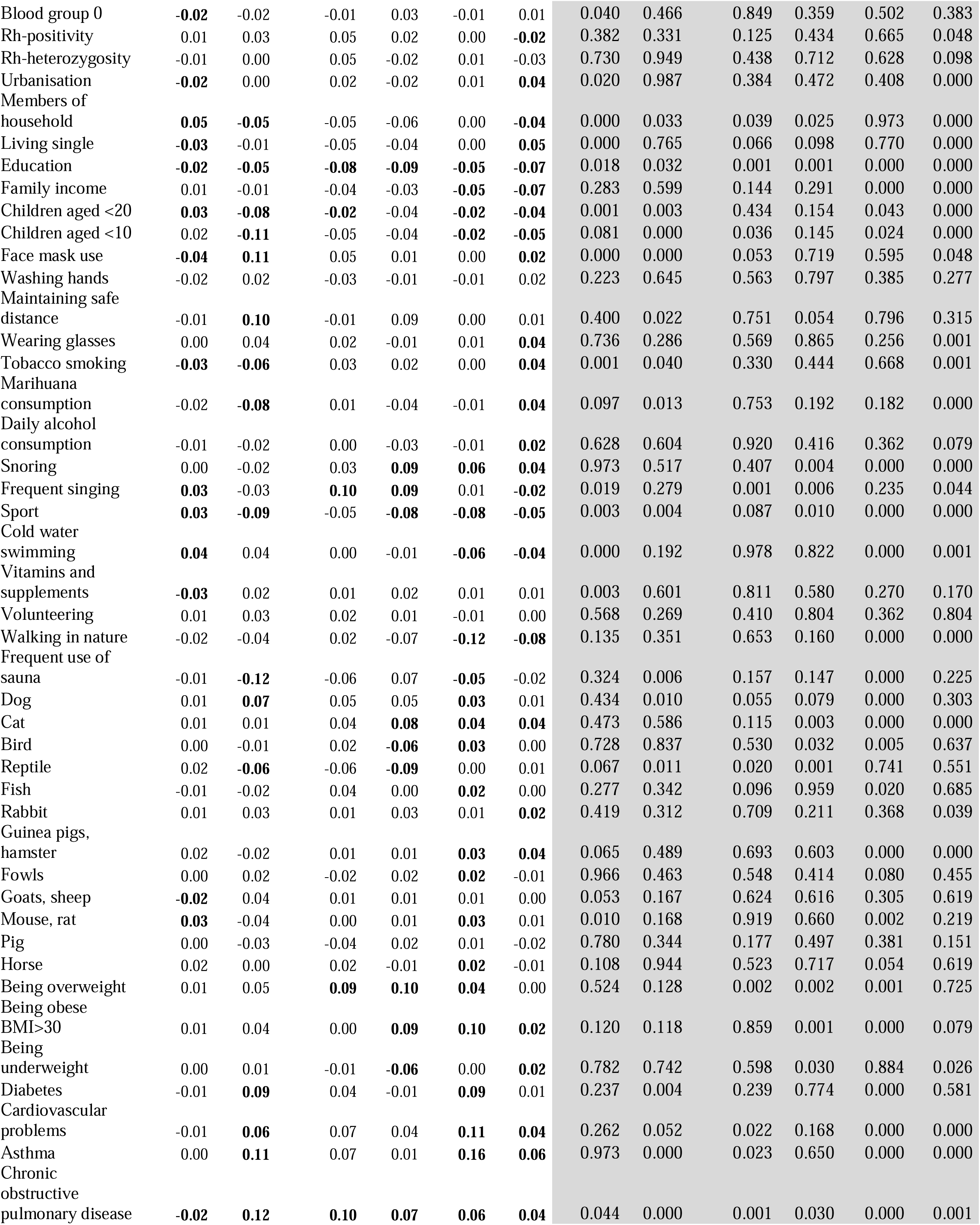
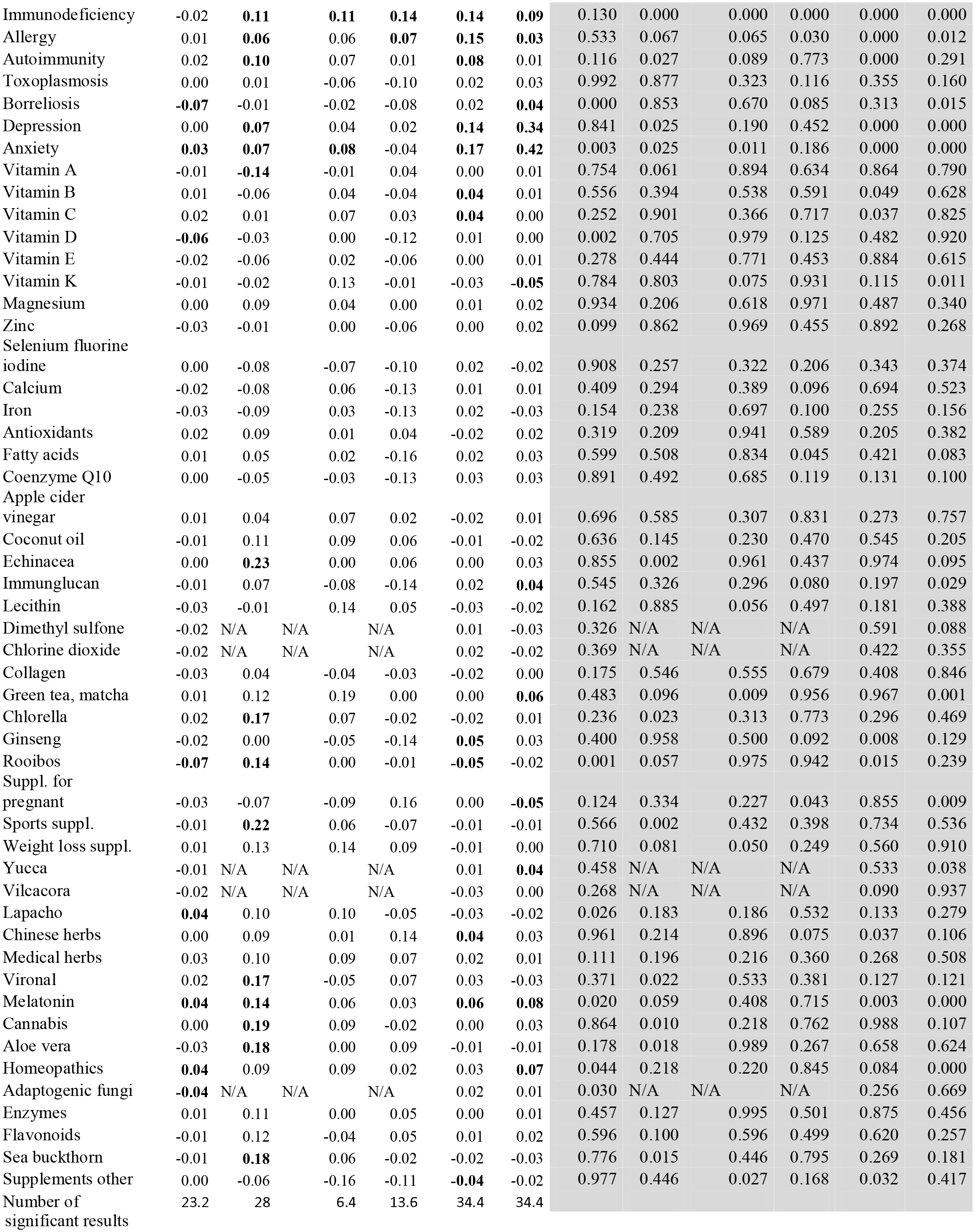
The effect of 105 factors on the risk of SARS-CoV-2 infection, Covid-19 course severity, and physical and mental health after the end of fourth wave of Covid-19 Columns 2–7 show the direction and strength (partial Kendall Tau) and columns 8–13 statistical significance of effects of the factors listed in the first column on the risk of Covid-19 infection (col. 2, 8), the course of Covid-19 (col. 3, 9), the severity of symptoms index (col. 4, 10), length of Covid-19 disease (col. 5, 11), and health after the fourth wave of Covid-19 – indices of physical and mental illness (col. 6–7 and 12–13). Positive Tau means a positive association between the factor in the first column and the dependent variable listed in the heading of the column. Taus printed in bold indicate associations which are significant after correction for multiple tests with the Benjamini-Hochberg procedure with false discovery rate 0.20 (20% of significant results in each column are false discoveries – artifacts of multiple tests). The number of significant results (without the 20% of false significant results) is shown in the last row. N/A means not available (cannot be tested), p-values under 0.0005 were coded as 0.000.

**Fig. 1.**
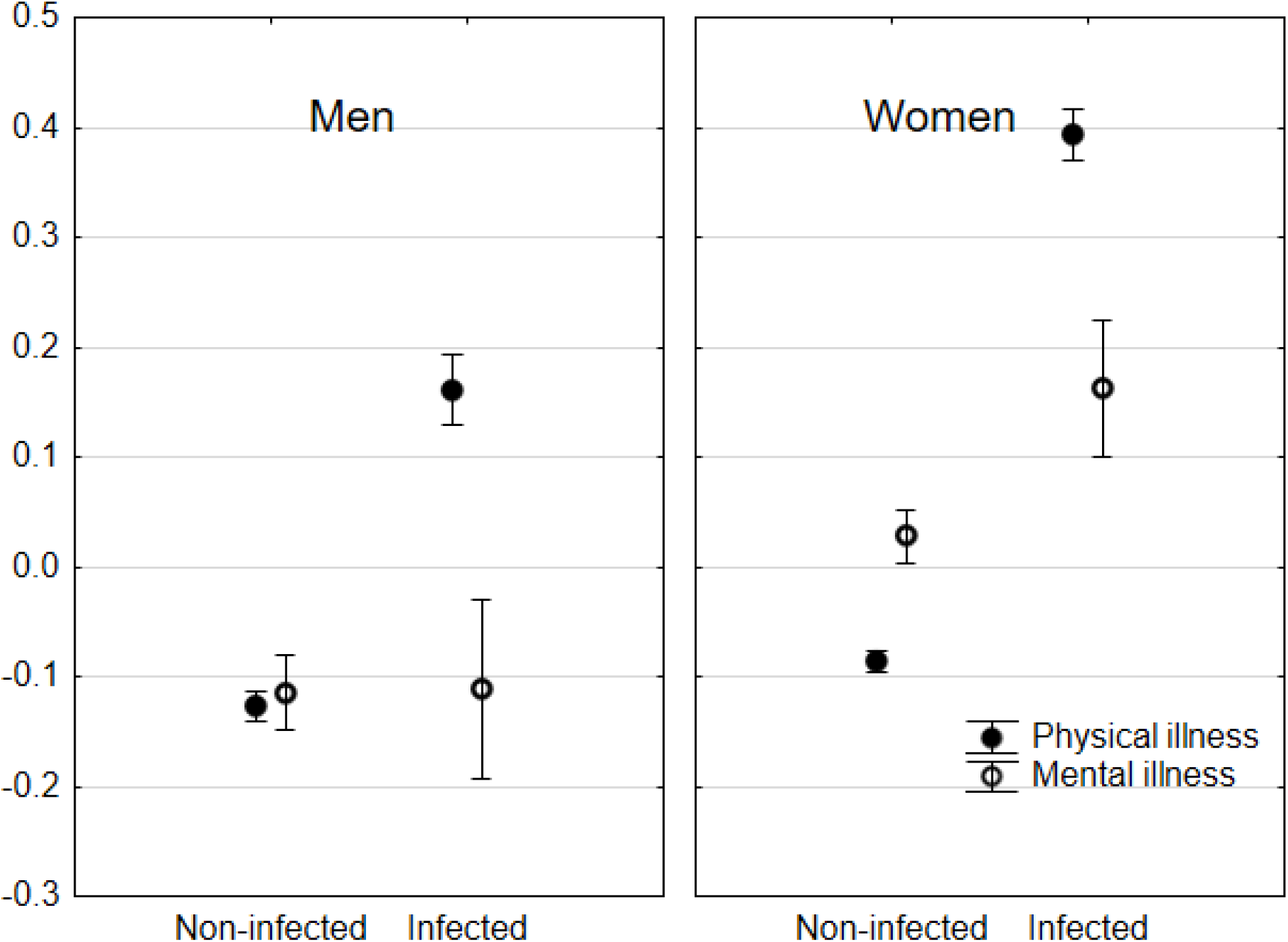
Mental and physical health of participants of the study after the end of the fourth wave of Covid-19 in the Czech Republic Numbers in graphs show the number of subjects in different categories; error bars show the 95% confidence interval.

To detect which biological, socioeconomic, behavioural, and environmental factors had a positive or negative effect on the risk of SARS-CoV-2 infection and risk of a severe course of Covid-19, we used separate partial Kendall correlation tests controlled for age, sex, and urbanisation level with 105 factors as independent factors, and variables infection with SARS-CoV-2 (yes/no), course of the Covid-19 infection (ordinal), severity of symptoms index (continuous), length of infection, and physical and mental health indices (continuous) as dependent variables. When age, sex, or urbanisation level was the subject of the analysis, only the other two remaining covariates were controlled for. Results of the analyses is shown in Table 3; results of analogical tests performed separately for each sex are displayed in Table 4.

**Table 4.** Effect of various factors on the risk of SARS-CoV-2 infection, severity of course of Covid-19, and post-Covid physical and mental health in women and men after the end of fourth wave of Covid-19 Results of partial Kendall analyses performed separately for women and men. For further information, see the legend of Table 3.

|  | Women |  |  |  |  |  | Men |  |  |  |  |  |
| --- | --- | --- | --- | --- | --- | --- | --- | --- | --- | --- | --- | --- |
|  | Infected | Course | Symptoms | Length | Physical illness | Mental illness | Infected | Course | Symptoms | Length | Physical illness | Mental illness |
|  | N/A | N/A | N/A | N/A | **0.27** | **0.06** | N/A | N/A | N/A | N/A | **0.23** | 0.00 |
| Age | -0.04 | **0.15** | 0.03 | **0.12** | **-0.05** | **-0.06** | -0.02 | **0.15** | -0.03 | 0.09 | **-0.03** | **-0.13** |
| Body height | 0.01 | 0.01 | -0.01 | 0.00 | -0.01 | **-0.02** | **0.05** | -0.03 | -0.01 | -0.04 | 0.00 | -0.01 |
| Body weight | 0.01 | **0.08** | 0.03 | **0.13** | **0.08** | 0.01 | **0.04** | 0.01 | 0.04 | 0.09 | **0.05** | -0.01 |
| BMI | 0.01 | **0.08** | 0.04 | **0.13** | **0.08** | **0.02** | 0.01 | 0.03 | 0.04 | **0.12** | **0.06** | -0.01 |
| Hair darkness | -0.04 | **-0.07** | -0.07 | -0.05 | **-0.03** | -0.01 | -0.01 | -0.01 | 0.00 | 0.03 | -0.01 | 0.01 |
| Hair redness | -0.01 | 0.00 | 0.03 | 0.04 | **0.03** | 0.01 | -0.02 | -0.05 | 0.01 | 0.03 | **0.05** | 0.03 |
| Skin darkness | -0.01 | **-0.09** | -0.05 | -0.03 | **-0.07** | **-0.05** | 0.01 | -0.03 | -0.03 | 0.02 | -0.02 | -0.02 |
| Blood group A | 0.00 | -0.03 | 0.01 | -0.03 | 0.00 | -0.02 | -0.01 | -0.03 | 0.01 | -0.06 | 0.01 | 0.02 |
| Blood group B | 0.02 | **0.08** | -0.01 | 0.01 | -0.01 | -0.01 | **0.06** | -0.04 | -0.06 | -0.02 | **0.04** | 0.00 |
| Blood group AB | 0.00 | 0.02 | 0.06 | 0.04 | 0.00 | 0.01 | -0.01 | 0.02 | -0.05 | -0.05 | 0.01 | -0.01 |
| Blood group 0 | -0.01 | -0.06 | -0.03 | 0.00 | 0.01 | 0.02 | -0.04 | 0.07 | 0.08 | 0.12 | **-0.05** | -0.02 |
| Rh-positivity | 0.02 | 0.05 | 0.03 | 0.02 | -0.01 | -0.02 | -0.03 | 0.01 | 0.12 | 0.06 | 0.00 | -0.03 |
| Rh-heterozygosity | -0.01 | 0.07 | 0.08 | 0.01 | 0.00 | -0.02 | 0.00 | -0.10 | -0.05 | -0.10 | 0.03 | -0.08 |
| Urbanisation | -0.02 | -0.05 | -0.01 | -0.06 | 0.01 | **0.04** | -0.03 | 0.08 | 0.08 | 0.06 | 0.01 | **0.05** |
| Members of household | **0.05** | **-0.10** | **-0.10** | **-0.11** | -0.01 | **-0.04** | 0.04 | 0.04 | 0.06 | 0.03 | 0.02 | -0.03 |
| Living single | **-0.04** | 0.02 | -0.02 | 0.00 | 0.02 | **0.06** | -0.03 | -0.05 | **-0.11** | **-0.12** | **-0.03** | **0.04** |
| Education | **-0.03** | **-0.11** | -0.07 | -0.07 | **-0.06** | **-0.07** | 0.00 | 0.02 | **-0.11** | **-0.12** | **-0.05** | **-0.05** |
| Family income | 0.00 | -0.03 | -0.04 | -0.04 | **-0.06** | **-0.07** | 0.03 | 0.02 | -0.03 | -0.01 | **-0.04** | **-0.07** |
| Children aged <20 | 0.02 | **-0.11** | -0.04 | -0.06 | **-0.02** | **-0.04** | **0.04** | -0.01 | 0.03 | 0.01 | -0.01 | **-0.04** |
| Children aged <10 | 0.01 | **-0.13** | **-0.10** | -0.06 | **-0.03** | **-0.04** | 0.03 | -0.07 | 0.03 | 0.00 | -0.01 | **-0.04** |
| Face mask use | **-0.03** | **0.11** | 0.05 | 0.05 | 0.01 | 0.01 | **-0.06** | **0.11** | 0.06 | -0.07 | -0.01 | 0.03 |
| Washing hands | -0.01 | 0.00 | 0.00 | -0.04 | 0.00 | 0.02 | -0.05 | 0.06 | -0.07 | 0.04 | -0.03 | 0.00 |
| Maintaining safe | 0.00 | **0.13** | 0.05 | **0.13** | 0.02 | 0.02 | -0.05 | 0.03 | -0.14 | -0.04 | -0.03 | -0.02 |
| distance |  |  |  |  |  |  |  |  |  |  |  |  |
| Wearing glasses | 0.01 | 0.04 | 0.02 | 0.04 | 0.02 | **0.05** | -0.01 | 0.02 | 0.00 | -0.13 | -0.01 | 0.01 |
| Tobacco smoking | **-0.03** | -0.03 | 0.05 | 0.04 | 0.02 | **0.04** | **-0.05** | -0.13 | -0.01 | -0.01 | **-0.05** | 0.02 |
| Marihuana consumption | -0.03 | **-0.08** | -0.07 | -0.04 | -0.02 | **0.04** | -0.01 | -0.08 | 0.06 | -0.06 | -0.01 | **0.05** |
| Daily alcohol consumption | -0.02 | 0.00 | 0.01 | -0.03 | -0.02 | **0.03** | 0.02 | -0.03 | 0.00 | -0.02 | 0.00 | 0.01 |
| Snoring | 0.01 | 0.04 | 0.04 | **0.14** | **0.07** | **0.06** | -0.02 | -0.10 | 0.02 | 0.02 | **0.04** | 0.03 |
| Frequent singing | 0.01 | -0.02 | **0.12** | **0.11** | -0.01 | **-0.04** | **0.06** | -0.06 | 0.06 | 0.05 | **0.06** | 0.03 |
| Sport | 0.02 | **-0.07** | -0.04 | -0.07 | **-0.08** | **-0.04** | **0.06** | -0.12 | -0.08 | -0.12 | **-0.09** | **-0.07** |
| Cold water swimming | **0.03** | 0.05 | 0.03 | 0.06 | **-0.04** | **-0.03** | **0.05** | 0.02 | -0.06 | -0.12 | **-0.08** | **-0.05** |
| Vitamins and supplements | **-0.03** | 0.02 | 0.01 | 0.02 | 0.02 | 0.00 | -0.03 | 0.00 | 0.01 | 0.01 | 0.00 | **0.05** |
| Volunteering | -0.01 | 0.02 | 0.03 | -0.03 | -0.02 | 0.00 | **0.04** | 0.07 | 0.04 | 0.07 | 0.00 | 0.00 |
| Walking in nature | -0.03 | **-0.12** | 0.00 | -0.07 | **-0.14** | **-0.08** | 0.01 | 0.15 | 0.09 | -0.05 | **-0.07** | **-0.09** |
| Frequent use of sauna | -0.01 | **-0.16** | -0.12 | 0.11 | **-0.05** | -0.02 | -0.02 | -0.07 | 0.05 | -0.04 | -0.04 | -0.01 |
| Dog | 0.01 | **0.09** | **0.08** | 0.07 | **0.02** | 0.00 | 0.01 | 0.02 | -0.01 | 0.01 | **0.06** | **0.04** |
| Cat | -0.01 | -0.01 | 0.05 | 0.05 | **0.03** | **0.05** | 0.03 | 0.06 | 0.03 | **0.13** | **0.05** | 0.02 |
| Bird | -0.01 | 0.01 | 0.01 | -0.04 | **0.03** | 0.01 | 0.02 | -0.04 | 0.01 | -0.09 | 0.02 | -0.01 |
| Reptile | 0.02 | **-0.08** | -0.05 | **-0.10** | 0.00 | -0.01 | 0.01 | -0.04 | -0.07 | -0.07 | 0.01 | 0.03 |
| Fish | -0.02 | 0.00 | 0.06 | 0.01 | **0.02** | 0.00 | 0.02 | -0.07 | 0.00 | -0.02 | 0.02 | 0.01 |
| Rabbit | -0.02 | 0.03 | 0.02 | 0.04 | 0.01 | **0.02** | **0.06** | 0.02 | -0.02 | 0.02 | 0.00 | 0.02 |
| Guinea pigs, hamster | 0.01 | 0.03 | 0.01 | 0.00 | **0.04** | **0.05** | 0.02 | **-0.10** | 0.01 | 0.05 | 0.03 | 0.02 |
| Fowls | 0.01 | 0.01 | 0.00 | 0.03 | 0.01 | -0.01 | -0.02 | 0.04 | -0.05 | 0.00 | 0.02 | -0.01 |
| Goats, sheep | -0.02 | 0.02 | 0.04 | 0.02 | 0.01 | -0.01 | -0.01 | 0.05 | -0.05 | 0.00 | 0.01 | 0.01 |
| Mouse, rat | **0.04** | -0.01 | 0.03 | 0.03 | **0.05** | **0.03** | -0.01 | **-0.14** | -0.09 | -0.04 | -0.01 | -0.03 |
| Pig | -0.02 | -0.01 | -0.01 | 0.03 | 0.01 | -0.02 | **0.05** | -0.05 | -0.07 | 0.01 | 0.01 | -0.01 |
| Horse | 0.01 | 0.03 | 0.06 | 0.01 | 0.00 | -0.02 | 0.03 | -0.08 | -0.06 | -0.06 | **0.07** | **0.04** |
| Being overweight | 0.02 | **0.09** | **0.11** | **0.13** | **0.04** | 0.00 | -0.01 | -0.05 | 0.07 | 0.01 | 0.03 | -0.02 |
| Being obese |  |  |  |  |  |  |  |  |  |  |  |  |
| BMI>30 | 0.00 | 0.04 | -0.03 | **0.08** | **0.12** | **0.03** | **0.04** | 0.04 | 0.03 | **0.12** | **0.08** | -0.01 |
| Being underweight | -0.01 | 0.00 | -0.02 | -0.05 | 0.01 | **0.03** | 0.01 | 0.04 | 0.02 | -0.07 | -0.02 | -0.01 |
| Diabetes | -0.02 | 0.05 | 0.02 | -0.03 | **0.09** | 0.01 | 0.00 | **0.14** | 0.07 | 0.02 | **0.07** | 0.01 |
| Cardiovascular problems | -0.02 | 0.03 | 0.03 | 0.06 | **0.08** | **0.04** | 0.00 | 0.11 | **0.14** | 0.03 | **0.16** | **0.04** |
| Asthma | -0.01 | **0.11** | 0.00 | -0.03 | **0.14** | **0.07** | 0.03 | 0.12 | **0.18** | 0.11 | **0.18** | 0.03 |
| Chronic obstructive pulmonary disease | -0.02 | **0.10** | **0.10** | 0.06 | **0.05** | **0.03** | -0.03 | **0.15** | 0.11 | 0.08 | **0.08** | **0.05** |
| Immunodeficiency | **-0.03** | **0.08** | **0.10** | **0.16** | **0.16** | **0.10** | 0.03 | **0.17** | 0.13 | 0.12 | **0.11** | **0.06** |
| Allergy | 0.01 | 0.04 | 0.04 | **0.10** | **0.15** | 0.02 | 0.01 | 0.10 | 0.08 | -0.01 | **0.14** | **0.05** |
| Autoimmunity | 0.00 | 0.09 | 0.06 | 0.01 | **0.07** | 0.01 | **0.09** | 0.12 | 0.09 | 0.01 | **0.09** | 0.02 |
| Toxoplasmosis | -0.01 | -0.06 | -0.14 | **-0.17** | 0.01 | 0.03 | 0.05 | 0.20 | 0.30 | 0.10 | 0.05 | 0.03 |
| Borreliosis | **-0.06** | 0.03 | 0.00 | -0.03 | 0.03 | **0.05** | **-0.07** | -0.09 | -0.08 | -0.19 | -0.01 | 0.02 |
| Depression | 0.00 | **0.09** | 0.02 | -0.01 | **0.14** | **0.36** | 0.01 | 0.01 | 0.06 | 0.11 | **0.14** | **0.32** |
| Anxiety | **0.05** | **0.10** | **0.09** | -0.05 | **0.18** | **0.42** | -0.01 | -0.01 | 0.04 | -0.03 | **0.16** | **0.41** |
| Vitamin A | 0.02 | **-0.18** | -0.03 | 0.11 | 0.00 | -0.01 | -0.06 | -0.02 | 0.06 | -0.24 | -0.01 | 0.04 |
| Vitamin B | 0.02 | -0.01 | 0.13 | 0.06 | **0.07** | 0.00 | 0.00 | -0.18 | -0.12 | **-0.42** | -0.03 | 0.02 |
| Vitamin C | 0.04 | 0.12 | 0.16 | 0.15 | **0.08** | 0.01 | -0.02 | -0.18 | -0.12 | -0.29 | -0.04 | -0.01 |
| Vitamin D | **-0.06** | -0.04 | 0.00 | -0.17 | 0.01 | -0.03 | -0.05 | -0.01 | 0.03 | -0.04 | 0.02 | 0.05 |
| Vitamin E | 0.00 | -0.07 | 0.08 | 0.00 | 0.01 | -0.02 | -0.05 | -0.02 | -0.09 | -0.24 | -0.02 | 0.06 |
| Vitamin K | -0.01 | -0.07 | 0.09 | -0.01 | 0.00 | -0.04 | 0.01 | 0.10 | 0.24 | 0.02 | **-0.09** | **-0.08** |
| Magnesium | -0.02 | 0.10 | 0.08 | -0.04 | 0.03 | 0.02 | 0.03 | 0.09 | -0.02 | 0.11 | -0.02 | 0.02 |
| Zinc | -0.03 | -0.03 | -0.02 | -0.06 | 0.00 | 0.01 | -0.03 | 0.05 | 0.08 | -0.04 | -0.01 | 0.04 |
| Selenium fluorine | 0.02 | -0.10 | -0.02 | -0.12 | 0.03 | -0.01 | -0.06 | N/A | -0.20 | N/A | 0.00 | -0.04 |
| iodine |  |  |  |  |  |  |  |  |  |  |  |  |
| Calcium | 0.00 | -0.11 | 0.08 | -0.12 | 0.03 | 0.00 | -0.06 | -0.01 | -0.02 | -0.19 | -0.03 | 0.03 |
| Iron | -0.02 | **-0.18** | 0.03 | -0.18 | **0.05** | -0.04 | -0.05 | 0.16 | -0.07 | 0.07 | -0.04 | -0.01 |
| Antioxidants | 0.00 | 0.07 | -0.04 | 0.01 | -0.03 | 0.00 | 0.05 | 0.17 | 0.18 | 0.23 | -0.01 | 0.04 |
| Fatty acids | -0.02 | 0.09 | -0.02 | -0.08 | 0.02 | 0.03 | **0.09** | -0.03 | 0.05 | **-0.46** | 0.00 | 0.04 |
| Coenzyme Q10 | 0.00 | -0.15 | -0.05 | -0.19 | 0.04 | 0.01 | -0.02 | 0.16 | -0.07 | 0.07 | 0.01 | **0.08** |
| Apple cider vinegar | -0.01 | 0.16 | 0.16 | 0.02 | 0.00 | 0.04 | 0.04 | -0.13 | -0.04 | 0.03 | -0.06 | -0.06 |
| Coconut oil | -0.02 | 0.09 | -0.02 | -0.01 | 0.00 | -0.03 | 0.03 | 0.14 | 0.29 | 0.34 | -0.04 | -0.02 |
| Echinacea | -0.01 | **0.25** | 0.05 | 0.04 | 0.01 | **0.05** | 0.01 | 0.21 | -0.13 | 0.24 | -0.02 | -0.04 |
| Immunoglucan | 0.00 | 0.08 | -0.10 | -0.16 | **0.06** | **0.07** | -0.05 | N/A | N/A | N/A | **-0.07** | -0.04 |
| Lecithin | -0.04 | N/A | N/A | N/A | 0.02 | -0.01 | -0.01 | 0.02 | 0.31 | 0.17 | **-0.09** | -0.03 |
| Dimethyl sulfone | -0.01 | N/A | N/A | N/A | 0.04 | **-0.04** | -0.03 | N/A | N/A | N/A | -0.02 | -0.02 |
| Chlorine dioxide | -0.02 | N/A | N/A | N/A | **0.05** | 0.01 | -0.02 | N/A | N/A | N/A | -0.04 | -0.06 |
| Collagen | -0.04 | -0.06 | -0.04 | -0.09 | 0.00 | 0.01 | 0.00 | 0.22 | -0.02 | 0.07 | -0.05 | -0.02 |
| Green tea, matcha | 0.01 | 0.01 | 0.00 | -0.13 | -0.02 | **0.04** | 0.02 | 0.32 | **0.53** | 0.31 | 0.04 | **0.11** |
| Chlorella | 0.03 | 0.16 | 0.09 | -0.05 | -0.03 | 0.03 | 0.00 | 0.16 | -0.07 | 0.07 | 0.01 | -0.01 |
| Ginseng | -0.01 | 0.02 | -0.08 | -0.01 | 0.04 | 0.01 | -0.02 | -0.01 | -0.07 | -0.30 | **0.07** | 0.05 |
| Rooibos | **-0.06** | 0.12 | -0.03 | -0.02 | -0.03 | 0.01 | -0.07 | 0.25 | 0.13 | 0.08 | **-0.09** | **-0.08** |
| Suppl. for pregnant | -0.04 | -0.09 | -0.10 | 0.17 | -0.01 | -0.05 | N/A | N/A | N/A | N/A | N/A | N/A |
| Sports suppl. | -0.02 | **0.18** | 0.11 | -0.14 | 0.02 | -0.02 | 0.01 | 0.29 | -0.02 | -0.03 | -0.05 | 0.00 |
| Weight loss suppl. | 0.01 | 0.15 | 0.16 | 0.09 | -0.01 | 0.00 | -0.02 | N/A | N/A | N/A | 0.00 | 0.00 |
| Yucca | -0.02 | N/A | N/A | N/A | 0.02 | **0.05** | N/A | N/A | N/A | N/A | N/A | N/A |
| Vilcacora | -0.02 | N/A | N/A | N/A | -0.03 | 0.01 | -0.01 | N/A | N/A | N/A | -0.03 | -0.04 |
| Lapacho | **0.06** | 0.13 | 0.13 | -0.05 | -0.02 | -0.02 | -0.01 | N/A | N/A | N/A | **-0.07** | -0.04 |
| Chinese herbs | 0.02 | 0.11 | 0.00 | 0.15 | **0.04** | 0.02 | -0.04 | N/A | N/A | N/A | 0.03 | **0.07** |
| Medical herbs | 0.02 | 0.09 | 0.04 | 0.04 | 0.04 | 0.01 | 0.06 | 0.08 | 0.19 | 0.21 | -0.02 | 0.03 |
| Vironal | 0.03 | **0.20** | -0.07 | 0.08 | 0.04 | -0.02 | -0.03 | N/A | N/A | N/A | 0.01 | **-0.08** |
| Melatonin | 0.02 | 0.08 | -0.07 | -0.04 | **0.07** | **0.10** | **0.10** | 0.21 | 0.22 | 0.16 | 0.03 | 0.03 |
| Cannabis | 0.00 | **0.28** | 0.09 | 0.01 | 0.02 | **0.04** | -0.01 | -0.07 | 0.02 | -0.23 | -0.05 | 0.01 |
| Aloe vera | 0.00 | **0.23** | 0.01 | 0.11 | 0.02 | 0.01 | -0.07 | N/A | N/A | N/A | -0.05 | -0.03 |
| Homeopathics | 0.04 | 0.13 | 0.04 | 0.00 | 0.03 | **0.07** | 0.04 | 0.02 | 0.31 | 0.17 | 0.06 | 0.06 |
| Adaptogenic fungi | -0.04 | N/A | N/A | N/A | 0.04 | 0.02 | -0.03 | N/A | N/A | N/A | -0.02 | -0.02 |
| Enzymes | -0.02 | N/A | N/A | N/A | -0.01 | 0.01 | **0.12** | 0.16 | -0.07 | 0.07 | 0.03 | 0.03 |
| Flavonoids | 0.00 | 0.15 | -0.05 | 0.07 | 0.00 | 0.01 | -0.03 | N/A | N/A | N/A | 0.02 | 0.05 |
| Sea buckthorn | 0.01 | **0.31** | 0.10 | 0.04 | 0.01 | -0.02 | -0.04 | -0.20 | -0.07 | -0.26 | **-0.09** | -0.04 |
| Supplements other | 0.01 | 0.09 | -0.06 | -0.14 | **-0.08** | -0.04 | -0.01 | -0.33 | -0.31 | -0.11 | 0.03 | 0.02 |

## Discussion

In this prospective cohort study, we analysed the effects of 105 potential protective and risk factors related to the incidence and severity of Covid-19 disease. We compared the incidence of Covid-19 and its severity (based on three different criteria), and both physical and mental health at the moment of filling the second questionnaire in subjects who had and had not been exposed to 105 focal factors before the start of the fourth wave of the Covid-19 epidemy in the Czech Republic. All participants were members of the Covid-negative cohort of internet users who shared with us information about their exposure to risk factors and protective factors in an electronic questionnaire distributed before the beginning of the fourth wave of the epidemic, on average 125 days before completing the second questionnaire. We grouped the factors into five categories: (1) biological factors including morphological traits, (2) sociodemographic factors, (3) behavioural traits/lifestyle variables, (4) contacts with animals, (5) comorbidities, and (6) use of vitamins and supplements.

In the first category, that of biological factors, we detected effects of sex and age on the risk of SARS-CoV-2 infection. Women and older subjects had a lower risk of infection; the possible role of behavioural immunity is discussed below. On the other hand, they also reported a more severe course of Covid-19. Only the latter corresponded to previously published findings ^1^. In general, women reported worse physical and mental health at the end of the study than men did. In accordance with the clinical experience and several published studies ^2,5,6,11^, individuals with higher weight and higher BMI experienced a more severe course of the disease. Surprisingly, taller and heavier men also ran a higher risk of infection than lighter and shorter men. Height was primarily responsible for this association because the association between infection and height was stronger than the association of infection with weight or BMI (the latter showed no association). In women, we found no association between height and increased risk of infection.

We should bear in mind, though, that questions about body weight and height were included only in the second questionnaire and the findings may have been influenced by the disease rather than being a risk factor of it. This is naturally not an issue for body height, which could not well change due to Covid-19, but it could have negatively influenced the effect size of association between body weight and the risk of SARS-CoV-2 infection and, although less so, it may have had an effect on the severity of course of Covid-19. It is likely that Covid-19, especially in case of a severe disease, has a negative effect on a person’s weight, which means that the association between body weight or BMI and infection rate and severity of Covid-19 is probably stronger than suggested by the strength of correlations detected in our study.

The lower risk of the infection in men and older subjects was probably due to increased effort of people who considered themselves especially at risk to avoid possible sources of infection: we observed the same phenomenon (in the form of significant effects or trends) in subjects with other known risk factors, such as immunodeficiency or chronic obstructive pulmonary disease. Notable exceptions (higher probability of infection in risk populations) were autoimmunity and obesity (BMI > 30) in men, which had relatively strong positive effects on the risk of infection. One could speculate whether these (and possibly also other) factors actually had a positive effect on the risk of infection or whether simply by their effect on the course of infection they increased the likelihood of a symptomatic course of Covid-19 and therefore also of the probability of the infection being recognised and officially diagnosed.

It has been generally expected that vitamin D ought to protect against Covid-19 ^12^ and it is known that redhaired individuals can synthesise more vitamin D in conditions of lower intensity of UV radiation, that is, in the higher latitudes of temperate zones ^13^. We have therefore expected that the intensity of red colour of hair would negatively correlate with the risk of infection or severity of Covid-19. A negative association between taking vitamin D supplements and risk of SARS-CoV-2 infection was confirmed by our data (see below) but we found no significant association between the intensity of red colour of hair and the risk of infection or a severe course of Covid-19. We only confirmed an earlier reported observation that redhaired subjects have a higher index of physical disease ^14^. It is possible that the favourable effect of having red hair and associated effect on the synthesis of vitamin D and the adverse effect of redhaired phenotype on physical health cancel each other out.

Our data showed that dark-haired women but not men had a lower risk of SARS-CoV-2 infection and a less severe course of Covid-19. This higher resistance of dark-haired subjects is probably the result of generally better health of dark-haired individuals in the Czech population ^13,15^. It is thus telling that dark-haired subjects – and even more so subjects with darker skin tone – had also a less severe course of Covid-19 (though it was significant only in women) and reported better physical health in the second questionnaire. It should be noted that for historical reasons, Czech population is ethnically highly homogenous and consists nearly exclusively of white Caucasian persons. The questionnaire was in Czech, a difficult Slavic language understood only by Czech and Slovaks. It is thus very likely that only ethnic Europeans took part in the study.

Blood group (system ABO) had a moderate effect on the risk of Covid-19 infection and probably no effect on its course. Individuals with blood group 0 had a lower and those with blood group B a higher risk of infection. The former concurs with the majority of published findings ^16,17^. The higher risk of the infection in subjects with blood group B also agrees with published data, but a meta-analytic study showed that blood group A usually has a stronger effect on the risk of Covid-19 than blood group B does ^18^. Both effects were stronger and statistically significant in men, while in women they were weaker and nonsignificant. Men with blood group B reported worse physical health in the second questionnaire, while those with blood group 0 reported better physical but worse mental health.

Rh factor had no significant effect on the risk of infection. Rh-positivity had only nonsignificant effects on the severity of course of Covid-19 (significant for the severity of symptoms index in men) which concurs with previously published data ^18^. Similarly, Rh-heterozygosity had no significant effect on the risk or severity of Covid-19, but that could be at least in part due to the relatively low number of participants whose heterozygosity could be determined based on their Rh-phenotype and the Rh-phenotype of their parents. Our results indicate that potential effects of Rh factor on the risk and severity of Covid-19 do deserve further attention, but investigation of this phenomenon should be preferably based on DNA-genotyped populations because Rh-positive heterozygotes have better and Rh-positive homozygotes worse health than Rh-negative individuals ^19^.

Sociodemographic factors had a moderate effect on the risks of Covid-19. People who live in larger cities and individuals with higher education, especially women, had a lower risk of infection, which is in agreement with published data ^20^. Household size, and in men especially the number of children under 20 years of age, was associated with a higher risk of infection, which again agrees with published data ^3,4^. People living on their own had a much lower risk of infection than those who share household with someone else and singles also reported a less severe course of Covid-19. Both of these effects were highly significant. Education level and in women also household size had the strongest protective effects against a severe or long course of Covid-19. Family income before the beginning of the pandemic had no significant effect on the risk of infection or the course of Covid-19 disease. This contrasts with findings of another prospective study which found a twice higher risk of Covid-19 in low-income individuals ^20^. That study, however, took into account only hospitalised patients. Income was positively correlated with physical and mental health at the moment of filling in the second questionnaire. It should be born in mind, though, that in the Czech Republic, nearly all medical care except for non-essential dentistry procedures and medical drugs that have cheaper alternatives is paid for from mandatory medical insurance. On the other hand, it is likely that higher-income individuals invest more in disease prevention.

Many behavioural traits had protective effects against the infection while three factors, namely being actively involved in sport (in both men and women), frequent singing (only in men), and cold water swimming (in both men and women), increased the risk of infection. We can only speculate about the proximal reasons of these findings. It seems likely that these activities increase the risk of infection only indirectly, that is, by increasing the number of physical contacts with other people. It is, however, also possible that singing facilitates the transmission of the virus even directly. A large community-based cohort study performed on 387,109 UK citizens showed a positive effect of physical inactivity on the risk of Covid-19 but the study took into account only hospitalised patients and not the much numerous subjects without a severe course of Covid-19 ^21^. The negative effect of sport on the risk of hospitalisation thus probably reflects the negative effect of physical activity on the risk of severe Covid-19 (observed also in our study), rather than its negative effect on the risk of the SARS-CoV-2 infection.

The strongest protective factor against Covid-19 infection was strict adherence to wearing masks and respirators; this factor was stronger in men than in women. Based on the results of laboratory tests, it is usually supposed that the wearing of masks, and even more so respirators, protects individuals against infection with SARS-CoV-2 (and not only against transmitting the infection to other people). On the other hand, the results of a metanalytic study show that empirical evidence for this claim is relatively weak ^22^. To the best of our knowledge, there is no published prospective longitudinal study that examined the effects of wearing masks on the risk of Covid-19 or its severity.

The second strongest protective factor was the consumption of vitamins and supplements. Analyses performed separately for women and men had shown that the strongest protective factor in women was walking in nature, possibly an indication of a solitary activity of more introverted women, because in men, walking in nature was a risk factor, albeit a weak and nonsignificant one, rather than a protective factor. The strongest protective factor for men was adherence to wearing masks and respirators. Sustaining social distance and frequent washing hands had only a weak and non-significant effect in both men (p-values > 0.069) and women (p-values > 0.699).

We found that tobacco smoking (in both men and women) and partly also of marihuana use (in women) have a relatively strong protective effect against SARS-CoV-2 infection. Marihuana use, and less probably also tobacco smoking, could have also some protective effect against a severe course of Covid-19. Protective effects of tobacco smoking have been reported ^7^ and discussed ^23^ in some previous studies but most studies show adverse effects of smoking on the risk of a severe course of Covid-19 ^2,11,21,24,25^. Former smoking habit seems to have a three times stronger adverse effect than current smoking ^26^, which agrees with the results of a metanalytic study based on 233 studies ^7^. We have no explanation for the contradiction between our data and reported data except for a hypothetical publication bias: it is possible that authors and editors may be reluctant to publish results showing any positive effects of smoking. It should be mentioned, though, that in our study, smokers reported worse mental health and female smokers reported worse mental and physical health in the second questionnaire than non-smokers did.

The most unexpected result of this part of the study was the positive correlation between higher severity of the course of Covid-19 and adherence to wearing masks and respirators and to a lesser extent also with keeping social distance. We speculate that individuals with predisposition to a severe course of Covid-19, that is, mainly those who were overweight, suffered immunodeficiency, chronic obstructive pulmonary disease, or diabetes, put more effort into trying to avoid infection and more strictly adhered to recommendations concerning wearing masks and maintaining safe distance. At the same time, if they did become infected they had a more severe course of the disease than individuals without such risk factors. The strength of these associations was lower or non-existent when the intensity of symptoms or duration of Covid-19 were used as a measure of severity of Covid-19 (except for the rather strong association between maintaining safe distance and duration of Covid-19 in women) and it was much stronger when we used a self-rated severity of the course of Covid-19. It is also possible that subjects who did not adhere to recommendations concerning personal protection against Covid-19 were later more reluctant to admit that they had a serious course of the disease. Alternatively, one could also speculate that more anxious people followed existing recommendations concerning individual protection against Covid-19 more strictly but they also tended to have a more severe course of Covid-19 if they did become infected. On the other hand, the strength of all the associations remained approximately the same when we included in the model reported intensity of anxiety and depression (partial Tau: masks 0.105 vs 0.109; distance 0.107 vs 0.107).

Coldwater swimming had a positive effect on physical and mental health at the time of filling the second questionnaire but it also seemed to be associated with a nonsignificantly more severe course of Covid-19 in women. Better immunity of people who are involved in this activity, which is popular in the Czech Republic, could have a negative effect on the course of Covid-19, possibly by increasing the risk of interleukin storm. A more probable explanation, however, is that subjects involved in this activity rarely suffer from seasonal colds, the flu, and another infectious diseases (either due to the effect of this activity or because only resistant people could perform such activity) and therefore rated the course of their Covid-19 infection as more serious than other individuals would.

In contrast, frequent use of sauna not only had a positive effect on physical and mental health (i.e., negative effect on the illness indices) at the time of filling the second questionnaire but was also negatively associated with a severe course of Covid-19. Taking all participants together, active sport and frequent use of a sauna had a strong protective effect against a severe course of Covid-19, the effect of sport being stronger in men, the effect of using a sauna in the woman.

Keeping certain animals could be a risk factor for acquiring the SARS-CoV-2 infection and it could also affect the risk of a severe course of Covid-19. Having cats or dogs as pets had no effect on the risk of infection and mostly nonsignificant positive effects on the risk of a severe course of Covid-19. The significant positive associations between dog keeping and more severe symptoms of Covid-19 in women (Tau = 0.095, p = 0.003) and between cat keeping and duration of Covid-19 in men (Tau = 0.134, p = 0.003) deserve future attention, but both could be just artifacts of multiple tests (see below). Similarly, the relatively weak effects of keeping other animals (rodents and pigs) on the risk of a more severe course of Covid-19 were probably just artifacts of multiple tests. It must be, however, reminded that hamsters are susceptible to the SARS-CoV-2 infection ^27^.

Known health-related predispositions to a worse course and outcome of Covid-19 mostly yielded the anticipated effects. The most severe impact was observed for immunodeficiency, autoimmunity, and chronic obstructive pulmonary disease but relatively strong were also the effects of being overweight, cardiovascular problems, and diabetes. Surprisingly, we did not detect any effect of latent toxoplasmosis, which was reported to be the strongest risk factor for the SARS-CoV-2 infection and for a severe course of Covid-19 in a previous cross-sectional study ^9^. It is rather unlikely that this discrepancy between results is due to differences in the experimental design (prospective cohort study vs. cross-sectional study). More likely is that the difference in risk factors could be caused by differences between the biological properties of the standard variant of SARS-CoV-2, which was the agent of all Covid-19 disease during the second and third wave of Covid-19, and alpha mutant of SARS-CoV-2, which was the agent of most Covid-19 cases during the fourth wave in the Czech Republic, which was the subject of the present study. It is known that not only infectivity but also the clinical picture of infection differs between the earlier and the beta variants of SARS-CoV-2 ^28^.

Another surprising finding was a very strong protective effect which having undergone borreliosis had against the infection in both sexes and, though only in men, also against a severe course of the disease. This effect has not been observed in the previous cross-sectional study ^9^. One could speculate that the extracellular parasite *Borrelia* redirects immunoreactivity of the host from humoral to cellular immunity, which might provide some protection against SARS-CoV-2. Moreover, the immunoregulative activity of *Borrelia* could provide some protection against a cytokine storm. And last but not least, borreliosis affects the physical and mental health, and secondarily also the behaviour of chronically infected subjects, which could likewise affect the risk of acquiring the SARS-CoV-2 infection [29]. As mentioned above, the protective effects against Covid-19 infection were relatively strong and significant in both women (Tau = −0.065, p = 0.0006) and men (Tau = −0.075, p = 0.009), but they could be the result of an artifact of multiple tests. In many countries, including the Czech Republic, seroprevalence of borreliosis is rather high ^29^. In the present study, it was 36% in Covid-negative and 26% in Covid positive participants. The observed protective effects, which seem to be stronger in men than in women, therefore deserve utmost attention in future studies.

All factors known to increase the risk of a severe course of Covid-19, with the exception of being overweight, provided some protection against acquiring the infection in women, but the effects were in nearly all cases nonsignificant. We suspect that people belonging to at-risk groups try more intensively (and at least partly successfully) to avoid contracting the infection. On the other hand, we did not observe any protective effect of depression or anxiety against acquiring the infection: in fact, more anxious women had a higher risk of acquiring Covid-19 and both depression and anxiety positively correlated with a higher probability of a more severe course of Covid-19 in women. This suggests that neither depression nor anxiety act as efficient instruments of human behavioural immunity against Covid-19.

During the epidemic, it has been suggested that regular taking of certain vitamins might act as prevention against Covid-19. People who live in the Czech Republic have often insufficient intake or photosynthesis of vitamin D and regular use of vitamin D supplements was therefore recommended by physicians as useful prevention against Covid-19. In our study, vitamin D provided significant protection against acquiring SARS-CoV-2 infection. Rather unexpectedly, though, the strongest protective effect against the infection was found for drinking rooibos, which is at least in the Czech Republic not considered a medical herb and it has not been suggested that it could help in Covid-19 prevention. It is known that rooibos, which is a fermented extract from the leaves of *Aspalathus linearis*, has both antioxidant and anti-inflammatory activities. Both in vitro and in vivo studies show that two major active dihydrochalcones found in the rooibos suppress vascular inflammation induced by high glucose or lipopolysaccharide in human vein endothelial cells. In mice, they suppress vascular inflammation caused by a wide range of molecular mechanisms including the inhibition of inflammatory cytokines and oxidative stress ^30-34^. It has been suggested by the authors of the corresponding study (performed on laboratory rodents) that aquatic extracts from the rooibos, i.e., rooibos tea, could be used to modulate oxidative stress and suppress inflammatory response ^35^. Moreover, thanks to the absence of caffeine in rooibos, it could be useful for reducing oxidative stress especially in children ^36^. As far as we know, no data on the effects of rooibos or its biologically active components have been published yet: an inquiry for rooibos AND Covid resulted in zero hits at WOS, Pubmed, MedRxiv, and BioRxiv.

This study had a character of exploratory research. All factors we planned to analyse were preregistered before the start of data collection to avoid the danger of cherry-picking artifacts. Nevertheless, the number of factors we examined (105) was so large that artifacts of multiple tests could be easily responsible for many significant results. It is mostly considered unnecessary or even counterproductive to perform a correction for multiple tests in exploratory studies ^37^ but in the present study, we decided (and preregistered) to perform this correction. To this purpose, we used the Benjamini-Hochberg method with a false discovery rate preset to 0.2, which is also why only 80% (140) of the 175 results indicated in bold in Table 3 as significant are expected to be significant in reality. In this context, it should be noted that the value of p before or after the abovementioned correction cannot itself discriminate between truly significant and false significant associations. For a discussion of the theoretical background of the method, relation between FDR and p-value, and superiority of controlling FDR over other methods of elimination of multiple tests artifacts, kindly refer to ^38,39^.

We would also like to draw attention here to the existence of a phenomenon of p-value spillover, that is, the effect of presence of many significant effects in a subset of factors (e.g. a subset of behavioural variables) on another subset of factors in which only a few or no effects exist (e.g. the subset of variables related to keeping animals). After the Benjamini-Hochberg or sequential Bonferroni correction, some significant effects in the former group will turn out to be nonsignificant and some nonsignificant effects in the latter group will become apparently significant.

The difficulty of recognising what is the cause and what the effect, what is a direct and what an indirect effect of a factor, and especially which factors affect the output variable and which merely indicate the existence of another (possibly an unknown) factor affecting the output variable are all serious problems affecting observational epidemiological studies. Unlike cross-sectional studies, longitudinal studies could discriminate between some alternatives but even these studies are not omnipotent. For example, by applying the Bradford Hill temporality criterium ^40^, we can be sure that the negative association between wearing masks (or taking vitamins) and acquiring the SARS-CoV-2 infection is not caused by a higher willingness of those who already had Covid-19 to protect themselves against the infection (or to treat symptoms or aftereffects of Covid-19). But we cannot exclude the possibility that some subpopulation of people protects itself against the infection in many ways, including wearing face masks, and that some of these methods of protection (but not the wearing of face masks) have a strong protective effect against Covid-19. Similarly, the observed strong positive association between taking echinacea and a severe course of Covid-19 could be caused by certain health problems which the subjects try to treat by echinacea and which also later predispose the subjects to a worse course of Covid-19, that is, it is possible that the effect is due to a kind of protopathic bias ^41^. The issue of causality could only be definitively solved by an intervention study, that is, by randomly assigning participants of a double-blind experiment into two groups and supplying one group with drug and the other with a placebo. Naturally, such experiments cannot be performed so as to investigate factors which are expected to have adverse effects on the course of a disease in humans. Also, it is sometimes technically difficult to perform a double-blind or blind experiment with some protective factors, such as wearing face masks.

### Strengths and limitations of the study

The most important advantage of the present study is its prospective longitudinal nature, its preregistration, and the large number of participants involved.

The most serious limitation of the study is the fact that participants were self-selected and do not represent a typical sample of a general population. The use of nonrepresentative samples (i.e., samples with less variability than is found in general population) increases the likelihood of finding even weak significant effects if they in fact exist. On the other hand, this setup could also artificially increase or decrease the observed strength of detected effects (the amount of variability in an output variable explained by the factors under study) ^42^. In short, due to the specific composition of the population of study participants, we must be careful with generalisation of the findings.

The second problem is that ‘survivorship bias’ could affect the results of some tests: Subjects who experienced a very severe course of Covid-19 were probably less likely to participate in the second part of the study (less likely to fill the second questionnaire) and those who died due to Covid-19 could not participate at all. In the Czech Republic, case mortality rate during the third and fourth waves of Covid-19 was about 1.9 % but the mean age of participants of our study was 43 and mortality in that age group was much lower. A low number of participants who died during the study, if any, could thus hardly affect the results of analyses aimed at identifying the risk and protective factors against the infection. On the other hand, a higher dropout rate of those participants who suffered a more severe course of the infection could affect the results of tests aimed at risk and protective factors against a severe course of Covid-19. It is, for example, possible that a large part of subjects with a certain risk factor, for instance those with chronic obstructive pulmonary disease or those with toxoplasmosis, had such a severe course of Covid-19 that they mostly did not participate in the second part of the study. Along similar lines, a seemingly milder course of Covid-19 in subjects who did not strictly adhere to mask wearing could be due to a survivorship artifact. There is probably no way of eliminating this kind of bias in questionnaire studies.

The third limitation of the study is the relatively low number of subjects affected by some factors. All in all, this study is based on a large number of subjects but the number of those who met a particular risk or protective factor could be rather low. For example, the number of subjects who drank rooibos and were not infected with SARS-CoV-2 was 121 (10.8%), while just 3 participants (3.3 %) drank rooibos and were infected with SARS-CoV-2. The equivalent numbers for, e.g., using marihuana, keeping rabbits, or being infected with *Toxoplasma* were 54/4, 783/132, and 153/25, respectively (see Table 1). Technically, a low or imbalanced number of subjects in particular groups is not a problem. Partial Kendall test is in principle an exact test and can thus be used to analyse this type of data, but small sample sizes and imbalanced distribution of observations in particular categories increases the risk of Type-1 error, i.e., increases the risk of not finding an existing effect. Of course, neither a small sample size nor imbalanced distribution could result in Type-2 errors, i.e., in detecting non-existent effects (see the Monte-Carlo model in the Appendix of ^43^).

## Conclusions

The present preregistered longitudinal study performed on a large population of internet users confirmed that some recommended measures, such as wearing masks or taking vitamin D, indeed protected participants against SARS-CoV-2 infection or a severe course of Covid-19, while other factors, even those that have a generally positive effect on health, such as sport or swimming in cold water, increased the risk of SARS-CoV-2 infection. The explorative nature of the study also brought some unexpected findings: for instance, we found a strong protective effect of being diagnosed with borreliosis in the past or drinking rooibos. Although the observed effects were strong and remained highly significant even after correction for multiple tests, it will be necessary to confirm their existence in future independent studies.

## Material and Methods

Participants were recruited by a Facebook-based snowball method ^44^. Calls for participation in the first part of the study were published about 15 times on the Facebook page of Labbunnies – a 23,000-member group of Czech and Slovak nationals willing to participate in studies on evolutionary psychology and evolutionary parasitology and to help with recruiting further participants of such studies – and on the authors’ personal Facebook and Twitter accounts. The Qualtrics questionnaire used to gather data contained Facebook ‘share’ and ‘like’ buttons, so that participants could help recruit other participants by pressing these buttons. The buttons were pressed 12,000 times between 17 October 2020 and 3 March 2021. In total, we obtained data from 52,000 respondents. In the end, though, many subjects finished the questionnaire up to four times at different time points (which they indicated in the questionnaire); only the first record of a participant was included in this study. The final set contained data from about 30,000 respondents. The invitation as well as the informed consent form on the first page of the questionnaire contained only the most general information about the aims of the study and contents of the questionnaire. The participants were informed that the study would examine which factors affect the risk of catching the new coronavirus and severity of the course of Covid-19 disease and investigate people’s views regarding anti-epidemic measures. Participants were also informed that their participation is voluntary, that they can skip any questions they might find uncomfortable, and that they can terminate their participation at any point simply by closing the web page. Only subjects who consented to participate in the study by pressing the corresponding button were allowed to take the questionnaire. Respondents were not paid for their participation but after finishing the 20-minute questionnaire, they received information about the results of related studies. The study was anonymous but participants had the option of providing their e-mail addresses for the purpose of a future longitudinal study (about 42% did) or could ask for their data to be deleted after completing the questionnaire (about 2% did). Data collection was performed in accordance with all relevant guidelines and regulations and the project, including the method of obtaining informed consent with participation in this anonymous study from all participants, was approved by the Institutional Review Board of the Faculty of Science, Charles University (Komise pro práci s lidmi a lidským materiálem Přírodovědecké Fakulty Univerzity Karlovy) — No. 2020/25). This first part of the study, including the questionnaire, was preregistered at the Open Science Framework: https://doi.org/10.17605/OSF.IO/VWXJE.

At the end of the fourth wave of Covid-19 in the Czech Republic, on 15 March 2021, we sent an email with an individualised link to the second electronic questionnaire to 12,600 subjects who provided their email address for this purpose at the end of the first questionnaire. About one-third of these emails have not been opened by the addressee, probably because they ended in their Junk or Spam folders. After two runs of reminders, the second questionnaire was filled by 8,084 subjects. This part of the project, a longitudinal prospective study, was preregistered at Open Science Framework (DOI 10.17605/OSF.IO/M7UVD).

## Questionnaires

Both surveys were run on the Qualtrics platform. The first questionnaire, which ran between 17 October 2020 and 3 March 2021, consisted of three parts related to three different projects (Risk and protective factors, Opinions of the Czech public regarding anti-epidemic measures, and the effect of priming by studying graphs of Covid victims on opinions regarding anti-epidemic measures).

In the present study, only responses to questions related to Covid-19 risks and protective factors were inspected and analysed. Respondents were asked about their sex, age, household size (this variable was also used for the calculating the binary variable single/non-single), family income before the beginning of the epidemic, and size of their place of residence (scale 1–5, 0: under 1,000 inhabitants, 1: 1–5,000 inhabitants, 2: 5–50,000 inhabitants, 3: 50–100,000 inhabitants, 4: 100– 500,000 inhabitants, 5: over 500,000 inhabitants). Respondents indicated whether they had already contracted Covid-19 by choosing from five answers (1: ‘No’, 2: ‘Yes, I was diagnosed with it’, 3: ‘Yes, but I was not diagnosed with it’, 4: ‘I am awaiting the test results’, 5: ‘No, but I was in quarantine’). For purposes of the current study, answers 1 and 5 were coded as 0 (Covid-negative), answer 2 as 1 (Covid-positive), and answers 3 and 4 were coded as NA (data not available).

In the main part of the questionnaire, respondents were asked to check which potential risks and protective factors apply to them, including keeping animals, taking vitamins and supplements, and being diagnosed with certain disorders often viewed as predisposing to a more severe course of Covid-19; for a list of corresponding binary variables, see column 1 of Table 1. In another part of the questionnaire, respondents were asked how strictly they follow measures related to personal protection against the infection, such as wearing masks, washing hands, and maintaining physical distance from other people. They had to answer the following three questions: ‘Do you abide by the measures concerning mask wearing/washing and disinfecting hands/maintaining safe distance (not to approach, not to touch)’ by choosing from five answers, namely 1: ‘No (on principle)’, 2: ‘No (due to indolence)’, 3: ‘Yes, but not too strictly’, 4: ‘Yes, I really strive’, 5: ‘Yes, strictly, and I try to convince people in my vicinity to do the same.’ Respondents were also asked whether they had ever been tested in a laboratory for toxoplasmosis and/or borreliosis and if so, what the result of this test was (negative/positive-infected/ ‘I do not know, I am not sure’). Similarly, respondents were asked about their blood AB0 group (possible answers: A/B/AB/0/ ‘I do not know, I am not sure’) and Rh status (positive/negative/ ‘I do not know, I am not sure’). For identifying the subpopulation of Rh-positive heterozygotes, we also asked them about their parents’ Rh phenotype ^19^. For questions regarding toxoplasmosis, borreliosis, and blood group, the questionnaire was pre-set to indicate the third response ‘I do not know, I am not sure’ as a default.

### The second questionnaire

The second questionnaire, which was disseminated in March 2021, contained again a question about whether participants had already contracted Covid-19. Those who had been diagnosed with it were also asked to rate the severity of the course of the disease on a five-point scale (1: ‘No symptoms’, 2: ‘Like a mild flu’, 3: ‘Like a severe flu’, 4: ‘I was hospitalised’, 5: ‘I was treated at an ICU’). They also had to check which symptoms they experienced during the Covid-19 infection. For a list of corresponding binary variables, see column 1 of Table 3. These variables were used for computing the severity of symptoms index as the mean z-score of all 22 variables. Participants were also asked to provide the dates of the beginning and end of their illness: this information was used to calculate the duration of the disease.

In another part of the questionnaire, respondents answered questions about their current physical health. They indicated how often they suffer from headache, rhinitis, gastrointestinal problems (problems including nausea, vomiting, or diarrhoea), sore throat or cough, allergy, sleeping problems, urinary tract inflammation, fatigue, and viral or bacterial infection, using an 8-point scale (1 – Never, 2 – Less than once a year, 3 – Once a year, 4 – Twice a year, 5 – Four times a year, 6 – Once a month, 7 – Once a week, 8 – More often). They also indicated how many drugs prescribed by physicians (except for contraceptives and drugs for mental health problems) they use and were asked to list which health problems (possible aftereffects of Covid-19) they ‘suffer from currently’ (fever, cough, breathlessness, sore throat, headache, stomach pain, diarrhoea, chest pain or pressure on the chest, conjunctivitis, middle ear pain, loss of smell, loss of taste, skin rash, changes in skin pigmentation, problems speaking and walking, fatigue, sniffles, sinus inflammation, joint and muscle pain, other pains, other health problems); these binary variables were coded 0/1. Then they rated how they are feeling currently in terms of their physical health using a graphic scale 0–100 anchored with 0 – Very well and 100 – Very bad. The index of physical illness was calculated as a mean z-score from these 32 variables. Participants also rated whether they suffer from depression and anxiety (two binary variables) and how often they suffer from depression, anxiety, and auditory hallucinations using an 8-point scale (1 – Never, 2 – Less than once a year, 3 – Once a year, 4 – Twice a year, 5 – Four times a year, 6 – Once a month, 7 – Once a week, 8 – More often), and how many drugs for mental health problems prescribed by medical professionals they take. Finally, they were asked to rate how they are feeling today in terms of their mental health using a graphic scale 0–100 anchored with 0 – Very well and 100 – Very bad. The index of mental illness was calculated as a mean z-score from these seven variables. In another part of the questionnaire, participants rated the darkness of their hair, their skin, redness of their hair, and provided information about their weight and height. They also answered how many children younger than 10 years and younger than 20 years live with them in the same household.

## Statistical analyses

Statistical analyses were performed with the R v. 3.3.1 software ^45^. To compute partial Kendall correlation, contingency table tests, and t-tests, we used the Explorer package ^46^. Correction for multiple tests was done using the Benjamini-Hochberg procedure with false discovery rate pre-set to 0·20 ^47^. The dataset is available at public repository Figshare 10.6084/m9.figshare.16529184 ^48^.

## Data Availability

All data are available at Figshare 10.6084/m9.figshare.16529184

https://10.6084/m9.figshare.16529184

## Acknowledgments

We would like to thank Anna Pilátová, PhD. for her help with preparing the final version of the article. This research was funded by Czech Science Foundation, grant number 18-13692S.

## Author Contributions

Conceptualization, original draft preparation, funding acquisition, and supervision, J.F.; formal analysis, writing—review and editing, investigation J.F., L.P. J.P. All authors have read and agreed to the published version of the manuscript.

## Data Availability Statement

All data are available at Figshare 10.6084/m9.figshare.16529184 ^48^.

